# Autoencoder Imputation of Missing Heterogeneous Data for Alzheimer’s Disease Classification

**DOI:** 10.1101/2024.07.18.24310625

**Authors:** Namitha Thalekkara Haridas, Jose M. Sanchez-Bornot, Paula L. McClean, KongFatt Wong-Lin, Alzheimer’s Disease Neuroimaging Initiative (ADNI)

## Abstract

Accurate diagnosis of Alzheimer’s disease (AD) relies heavily on the availability of complete and reliable data. Yet, missingness of heterogeneous medical and clinical data are prevalent and pose significant challenges. Previous studies have explored various data imputation strategies and methods on heterogeneous data, but the evaluation of deep learning algorithms for imputing heterogeneous AD data is limited. In this study, we addressed this by investigating the efficacy of denoising autoencoder-based imputation of missing key features of a heterogeneous data that comprised tau-PET, MRI, cognitive and functional assessments, genotype, sociodemographic, and medical history. We focused on extreme (40-70%) missing at random of key features which depend on AD progression; we identified them as history of mother having AD, APoE ε4 alleles, and clinical dementia rating. Along with features selected using traditional feature selection methods, we included latent features extracted from the denoising autoencoder for subsequent classification. Using random forest classification with 10-fold cross-validation, we evaluated the AD predictive performance of imputed datasets and found robust classification performance, with accuracy of 79-85% and precision of 71-85% across different levels of missingness. Additionally, our results demonstrated high recall values for identifying individuals with AD, particularly in datasets with 40% missingness in key features. Further, our feature-selected dataset using feature selection methods, including autoencoder, demonstrated higher classification score than that of the original complete dataset. These results highlight the effectiveness and robustness of autoencoder in imputing crucial information for reliable AD prediction in AI-based clinical decision support systems.

## I. Introduction

Alzheimer’s disease (AD), the most common cause of dementia, is a progressive brain disorder associated with memory loss, affecting day to day activities and cognitive decline [1]. Detecting AD and its severity level at early stage can enable better disease management and reduced care costs [2]. Further, adopting the right measures in clinical diagnosis is essential for timely treatment, care and disease management. Several assessment strategies and markers are currently available, including brain/blood-based biological assessment, medical and family history, and neuropsychological assessments [3]. Due to the variety of assessments and with symptoms overlapping with normal ageing and other types of dementia [4], diagnosis of AD remains challenging.

The implementation of technology-aided decision support system, especially involving machine learning, for diagnosis may offer a promising path [5]. However, when clinical data contains missing values, it significantly impacts early diagnosis and treatment, underscoring the importance of implementing effective measures to uphold data quality and integrity [6]. In particular, missing data, if not handled appropriately, can potentially delay treatment and lead to biased diagnostic results, including in machine learning decisions.

Clinical data may be incomplete in different scenarios. For instance, patients may not arrive for medical appointments or being unable to complete surveys [6]. Incomplete data is highly prevalent in cohort studies, especially within dementia and AD studies due to factors such as longer study requirement, increased risk of mortality and cognitive decline among older adults. These factors impede their ability to participate in studies requiring multiple visits, leading to missing data [7]. More generally, missing data can be classified into three types: missing at random (MAR) that is dependent on observed variables, such as when the proportion of missingness increases with dementia severity; missing completely at random (MCAR), in which the missingness is independent of any variables; and missing not at random (MNAR), in which missingness is dependent on unrecorded variables [8]. The trivial solution is to ignore the missing portion, but that may lead to low statistical power for the machine learning models. Therefore, it is imperative to employ appropriate strategies for data imputation [6].

Previous studies have made use of different imputation methods on clinical datasets in different contexts [9], [10]. For example, in a study [11] that focused on the Alzheimer’s Disease Assessment Scale – Cognitive Subscale 13 (ADAS-Cog 13) within the ADNI dataset, it demonstrated the importance of imputation by analysing longitudinal ADAS-Cog 13 scores and their association with baseline patient characteristics. By employing multiple imputation by chained equations (MICE), the study showed that imputation led to valid model estimates with tighter confidence intervals, thereby improving the efficiency of statistical analysis. However, the study primarily focused on ADAS-Cog 13, which may limit the generalisability to more comprehensive data.

Another study [12] investigated the impact of common imputation methods on missing AD data, with 33% of missing data, primarily associated with PET imaging data. Instead of discarding incomplete data and analysing only complete data, they explored various imputation techniques to enhance the dataset. Following imputation, they trained support vector machine (SVM) and random forest classifiers to distinguish between different progressive levels of AD. Their findings underscored the significance of employing imputation procedures to enhance classification accuracy and robustness. Again, the study did not involve more comprehensive data.

Inspired by the missingness structure (MAR or MCAR) in real-world memory clinic data, a recent study [6] synthetically generated similar missingness in an open complete dataset. It systematically evaluated multiple imputation and AD classification workflows, having missingness in both the training and testing datasets for machine learning, and using the original complete data as ground truth for post-imputation evaluation. The study showed that iterative imputation on the training dataset combined with a reduced-feature classification model as the most effective approach in terms of speed and accuracy in the imputation. More generally, classification accuracy of progressive stages of AD did not vary widely across imputation and classification strategies, but the computational cost can be orders of magnitude different. However, the study was limited to cognitive and functional assessments (CFAs).

Another recent study systematically addressed the challenge of missing data in AD diagnosis by identifying the gradient boosting imputing algorithm to be the best for missing values AD dataset [13]. Although this study used more heterogeneous and comprehensive data than others, they did not involve sub-assessments of CFAs, the MRI data modality was limited to a small number of brain regions, and their PET data focused on metabolic (FDG) and beta amyloid plaques (PiB amyloid and AV45 amyloid). The tau pathology data was based on cerebrospinal fluid (CSF). However, it is known that brain tau pathology has superior diagnostic utility than brain beta amyloid pathology and CSF tau [14]. Moreover, deep learning was not evaluated and the authors made use of missingness of the MCAR type and not on key data features. The classification accuracy after imputation was rather low when missingness level was high; hence, the imputation might lack robustness.

So far, the imputation methods in the abovementioned studies have made use of traditional machine learning algorithms. Autoencoder, a deep learning technique, is increasingly used for imputing missing data due to their ability to learn efficient embeddings (low-dimensional latent features) of unlabelled data [15]. In fact, autoencoders have been shown to outperform traditional machine learning algorithms, e.g. k-nearest neighbors, mean, median, RawInput, and SoftInput [8].

A recent study [16], the only study that has used autoencoders to impute missing AD data, used baseline data consisting of mini mental state examination (MMSE), magnetic resonance imaging (MRI), positron emission tomography (PET) (with unspecified tracer), cerebrospinal fluid (CSF) data, and other personal information. Principal component analysis (PCA) was used for reducing the dimensionality of data while SVM was used for binary classification between classes control normal (CN), mild cognitive impairment (MCI, a mixed group which includes prodromal stage of AD), and AD. The personal information comprised gender, age and marital status, without family history of AD. Moreover, there was only one cognitive and functional assessment, the MMSE. Thus, the data was not sufficiently detailed and comprehensive. The type of autoencoders used was also unspecified. Importantly, missing data was not systematically evaluated in that study.

To address the limitations of previous studies on missing data imputation for AD classification, in the current study, we focused on the imputation of the missingness of the most important AD data features with respect to predicting different progressive stages of AD. These key data features were common features identified from multiple feature selection methods, including with an autoencoder to elucidate latent features. MAR missing data were then systematically generated over different proportion of missingness from a complete, comprehensive an open dataset, with the latter acting as ground truth. This was followed by the implementation of denoising autoencoder on the missing key features. Finally, 3-class classification (CN, MCI and AD) of the imputed datasets by the random forest classifier was employed, and the results were compared with those based on the classification of the original complete datasets and the subset of selected features.

## II. Methods

### A. Data Description

The Alzheimer’s Disease Neuroimaging Initiative (ADNI) dataset, more specifically the ADNIMERGE-3 open repository, was employed as the primary data source for this study. The data was obtained from the https://adni.loni.usc.edu portal as per request and was accessed after approval by the Data Sharing and Publication Committee of Image and Data Archive (IDA). The dataset comprises of clinical, neuropsychological assessments, neuroimaging measures, and genetic markers collected from participants diagnosed as healthy (CN), having mild cognitive impairment (MCI), or Alzheimer’s disease (AD). The processed neuroimaging (tau-PET and structural MRI) is available from a previous study [17]. The final processed dataset consists of 559 samples (participants) encompassing a total of 224 features.

The processed dataset comprised 7 sociodemographic and medical history features, 40 CFA scores including their sub-assessments, and a relatively extensive 177 neuroimaging features (from active tau-PET brain regions via co-registering with structural MRI [17]). The class labels for training the classification model were based on clinician diagnosis, comprising 363 CN, 137 MCI and 59 AD cases.

The sociodemographic and medical or family background features comprised of age, gender, years of education, maternal and paternal family history of AD, and the number of copies of the APoE ε4 alleles (henceforth referred simply as APoE4). The CFAs’ scores were derived from various measures, including the Alzheimer’s Disease Assessment Scale (ADAS), Cognitive Battery Assessment, Clinical Dementia Rating (CDR), Mini-Mental State Exam (MMSE), Modified Hachinski Ischemia Scale, Neuropsychological Battery Test, logical memory immediate recall test (LMIT), logical memory delayed recall test (LMDT), the Neuropsychological Inventory (NPI), and the Geriatric Depression Scale (GDS). The dataset included individual question scores from ADAS and individual subscales from NPI, while other CFAs and individual CFA subscales from ADNI were excluded due to significant missing data. The tau-PET neuroimaging data utilised the [^18^F]AV-1451 tracer for detecting tau deposition [18].

### B. Data Preparation and Preprocessing

As part of the preprocessing pipeline, the participant identification (ID) column (‘RID’) was first removed. The gender column (‘PTGENDER’) was standardised by subtracting 1 from its values to ensure consistency in representation [19].

Certain columns in the dataset contained negative values, which required adjustment to ensure compatibility with subsequent processing steps. The columns with negative values were identified, all of them were CFA columns including COMP_MEM_SCORE, PHC_MEM, PHC_EXF, PHC_LAN, and COMP_EXEC_FUNC_SCORE, then transformed by adding the absolute value of their minimum to all values, effectively shifting the distribution to non-negative values.

Afterwards, normalisation [20] of features was performed as a part of preprocessing using min-max scaling method [21]. Features with a maximum value exceeding 1 were identified, excluding the target variable (AD_LABEL) and clinical dementia rating (CDR). These features were then normalised by subtracting their minimum value and dividing by the range between the maximum and minimum values. The preprocessed dataset was then used for feature selection.

### C. Feature Selection

#### 1) Boruta

The Boruta algorithm [22] is a feature selection technique based on random forests, designed to distinguish relevant features by comparing their importance with that of random features. It iteratively evaluates the significance of each feature and selects those that demonstrate significance above a predefined threshold. In this study, the Boruta algorithm was chosen for its ability to handle complex datasets with high-dimensional features. Moreover, Boruta harnesses the collective learning strengths of random forests, ensuring robust feature selection by considering feature interdependencies [22].

#### 2) Logistic Regression with L1 Regularization (Lasso)

Lasso regression with L1 regularisation [23], also known as Lasso regression, is a linear model which penalises the absolute magnitude of the coefficients, produces sparse solutions, with certain coefficients being set to zero. This property makes Lasso regression suitable for feature selection by identifying and prioritising the most relevant features while disregarding irrelevant ones [24]. In this study, Lasso regression was employed to complement the Boruta algorithm and provide additional insights into feature importance based on the model coefficients.

#### 3) Autoencoder

Autoencoder-based feature selection implements deep learning techniques to learn the compact representations of high-dimensional data. By training an autoencoder model to reconstruct the input features, the encoder layer learns to capture the essential information from the data, effectively performing feature extraction [25]. In this approach, a three-layer autoencoder was utilized, comprising an input layer, an encoding layer, and a decoding layer. The input layer defined the shape of the input data, while the encoding layer, implemented as a dense layer with encoding neurons and ReLU activation, learns to encode the input features into a lower-dimensional representation, capturing the most important features. Subsequently, the decoding layer, composed of a dense layer with the same number of neurons as the input layer. This autoencoder-based feature selection was employed to unveil the latent features that contribute significantly to the variability in the ADNI dataset. Notably, this approach complements traditional feature selection techniques by capturing nonlinear relationships and revealing hidden patterns in the data.

### D. Generating Missing Data of Selected Features

MAR is a common issue in AD data, influencing the reliability of analyses and the effectiveness of predictive models [26]. To simulate MAR, we intentionally introduced varying degrees of missingness (40%, 50%, 60%, and 70%) individually in the three pivotal variables of our dataset based on the selected key features that were more likely to be missing with later progressive stage of AD [6]. For example, we first identified the maternal dementia history (MOTHDEM) column as a key feature for MAR generation. Then, to introduce specific level of missingness in the MOTHDEM column, we devised a method that relies on the relationship between MOTHDEM and another pivotal variable, such as CDR, that is objectively and strongly associated with AD diagnosis. Specifically, this method introduces missingness in MOTHDEM based on CDR values. For example, if CDR indicates MCI (CDR = 0.5), we randomly select a fraction of observations and make the corresponding MOTHDEM values missing; higher missingness fraction for AD (CDR = 1). This approach allows us to simulate missingness patterns in MOTHDEM that depend on the progressive stage of AD indicated by CDR, thereby creating a more realistic representation of MAR scenarios [6]. For missing CDR, we tagged missingness levels with AD diagnosis.

### E. Imputation using Denoising Autoencoder

Unlike traditional imputation techniques that depend solely on statistical methods or simple interpolations, denoising autoencoders harness the capabilities of deep learning architectures to learn the intricate patterns and relationships present in the data [27]. The architecture of a denoising autoencoder comprises three main components: an encoder, a decoder, and a corruption process. The encoder reduces the dimensionality of the input data, mapping it to a latent space representation. The corruption process introduces noise or distortions to the input data to make the autoencoder to learn robust features. The decoder then reconstructs the original input from the compressed representation [28].

During training, denoising autoencoders strive to minimise the reconstruction error between the input and its corresponding reconstruction, typically employing mean squared error as the optimisation metric. This iterative process enables the model to adeptly learn meaningful representations of the data by denoising and reconstructing inputs, rendering them invaluable for data denoising tasks. By training the autoencoder on the observed data while deliberately introducing noise or corruption, the model becomes proficient at reconstructing the original, noise-free data. Consequently, denoising autoencoders excel in effectively filling in missing values through the learned representations [29].

In this study, after synthetically generating varying proportion of missingness intentionally in the columns of the selected key features of the dataset, a denoising autoencoder was employed for all imputations. The architecture of the denoising autoencoder comprised an input layer with dimensions equivalent to the number of input features in the dataset, followed by two hidden layers with 126 and 63 neurons, respectively. Dropout layers with a dropout rate of 0.2 were strategically incorporated after each hidden layer to mitigate overfitting and enhance the model’s robustness. Rectified linear unit (ReLU) activation functions [30] were utilised in the hidden layers to capture nonlinear relationships within the data effectively. Gaussian noise [31] with a standard deviation of 0.1 was introduced to the input layer to augment the denoising capability of the autoencoder, facilitating the accurate reconstruction of missing values while mitigating noise. The output layer employed linear activation to ensure continuous output values.

The autoencoder was trained using the mean squared error (MSE) loss function [32] and the Adam optimiser [33] over 50 epochs [34], with a batch size of 50 and a validation split of 20%. Then the autoencoder was able to predict and impute missing values in the respective columns, thus contributing to the restoration of data integrity and enhancing the reliability of subsequent analyses.

Following the imputation process using the denoising autoencoder, the performance of the imputation was evaluated using two key metrics: root mean squared error (RMSE) [35] and imputation accuracy [36]. RMSE was computed to quantify the discrepancy between the actual values of the missing column and the imputed values generated by the autoencoder. This metric provides a measure of the average difference between the observed and predicted values, thereby assessing the imputation accuracy on a continuous scale. Additionally, imputation accuracy was calculated to assess the correctness of the imputed values produced by the autoencoder. The imputation accuracy was computed as the ratio of the number of correctly imputed values to the total number of imputed values, providing a percentage value indicative of the accuracy of the imputation process.

Following the imputation process, a visual comparison between the actual and imputed values of the selected columns were conducted using a scatter plot. Additionally, the feature importance of the denoising autoencoder was examined by analysing the weights of the first dense layer. This analysis allowed the identification of the most influential features for accurate imputation, contributing to a better understanding of the model’s performance.

### F. Classification and Model Performance Evaluation

In the classification phase of the work, random forest classifier [37] was employed due to its capability to handle complex data and mitigate overfitting. It also performs especially well in heterogeneous AD data [38]. This ensemble learning technique combines multiple decision trees, providing robustness to noisy data and reducing the risk of overfitting.

We employed 3-class classification for classifying the 3 classes of the target variable AD-LABEL, namely, the CN, MCI and AD groups. Additionally, imbalance observed in the classes was handled by utilising the synthetic minority over-sampling technique (SMOTE) [39] and ensured that the classifier could effectively learn from all classes.

Evaluation of the random forest classifier was performed using 10-fold cross-validation [40], a robust method for estimating the model’s performance on unseen data. The classifier was trained on resampled training datasets to mitigate the impact of biased sampling, and its performance was assessed using the following metrics: accuracy, precision, recall and F1-score [41]. Additionally, the confusion matrix [42] and classification report were generated to provide detailed insights into the classifier’s predictive performance for each class label.

We conducted classification using random forest classifier for all imputed datasets as well as the original complete dataset. Additionally, we evaluated the performance of classification models using features selected from three different feature selection methods: Boruta, logistic regression, and autoencoder. Furthermore, we included features extracted from the denoising autoencoder as part of our analysis. The overall process of the current study is schematically shown in Fig 1.

**Fig 1.**
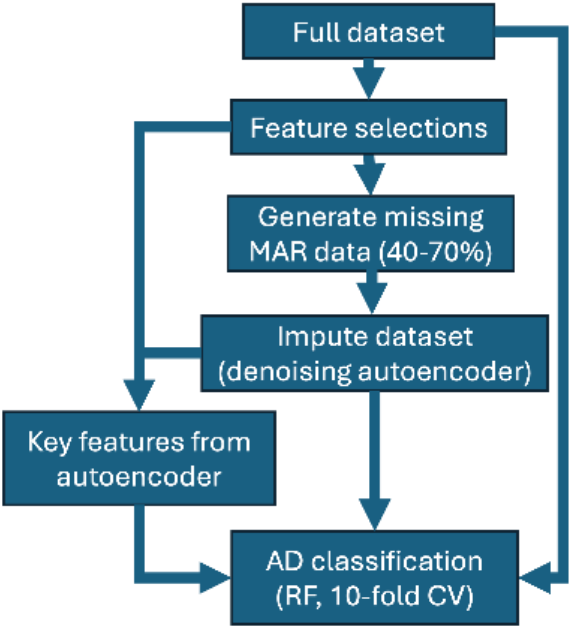
Schematic of workflow of current study.

### G. Software

Google Colab (https://colab.google/), a cloud-based Jupyter notebook environment provided by Google, was utilised for conducting the study, which included data preprocessing, feature selection, missing data generation, imputation, classification and validation tasks. Codes are available upon reasonable request.

## III. RESULTS

### A. MAR of consistently extracted features

Feature extractions were performed using Boruta, logistic regression with L1 regularization and autoencoder with respect to AD severity and the common top-ranked features were extracted. Amongst the data comprising demographic, CFAs, genetic, and neuroimaging markers, we identified maternal dementia history (MOTHDEM) and gender as influential factors, with maternal dementia history serving as a potential genetic predisposition and gender reflecting sex-related differences in AD risk. Further, the key neuroimaging (region-of-interest, ROI) data features identified were the left hemispheric amygdala, white matter left hemispheric entorhinal and inferiortemporal cortices, consistent with known neurodegenerative changes associated with AD progression [43]. Additionally, the genetic biomarker APoE4 were identified, highlighting its significant role as an AD risk factor, and the clinical dementia rating (CDR), which has been shown to be closely correlated with AD severity [44].

Amongst these extracted features, MOTHDEM, APoE4 and CDR are more likely to be affected by AD severity. For instance, with more severe AD, the patients may not remember their mother had AD. Patients with more severe AD may also be less likely to undergo genetic screening, which require specialised technical facility. As CDR takes about 90 minutes to administer [45], relatively long amongst the considered CFAs, it is more probable that patients with severe AD will not have the patience or ability to undergo such long assessment. Although it is plausible that patients with overt AD may also be less likely to undergo PET scans, the processed dataset was dominated by PET-MRI’s active ROIs and it is in practice unlikely that individual imaging ROI features would be missing. Hence, we did not remove any PET-MRI features in this study. Nevertheless, even without this, our proposed columnar missingness could incur substantial reduction in classification accuracy (see below).

For the MOTHDEM and APoE4 variables, missing data of MAR type were generated based on the clinical dementia rating (CDR) values, on the samples (rows) with CDR values of 0.5 or 1 (the higher the value, the more severe the condition) at the probability of missingness (0.4, 0.5, 0.60, 0.7) Similarly, for the CDR variable, missing data of MAR type was introduced, but this time it was based on the clinical diagnosis (AD_LABEL), where samples (rows) labelled as MCI or AD were more likely to have missing CDR values at the probability of missingness (0.4, 0.50, 0.6, 0.7). See Section II.D for further details. Although we focused on only the missingness of these three variables, which had strong influence on detecting AD severity (via feature selection), we generated large proportions of missingness within these variables (from 40% to 70% missing data per variable) and classification was substantially poorer (see below).

### B. Imputation with denoising autoencoder

Next, denoising autoencoder was employed separately to perform the imputation of missingness introduced 40%, 50%, 60% and 70% for the pivotal columns MOTHDEM, APoE4 and CDR. Further to visualise the accuracy and reliability of our imputation, we plotted scatter plots comparing the actual and imputed values and showed their similarities (Supplementary Fig. 1). To gain deeper insights, we extracted important features from the dense layer of the autoencoders by calculating the feature importance from each imputed dataset. Table I shows the full list of the commonly selected features using the employed traditional feature selection methods and denoising autoencoders.

**TABLE I.**
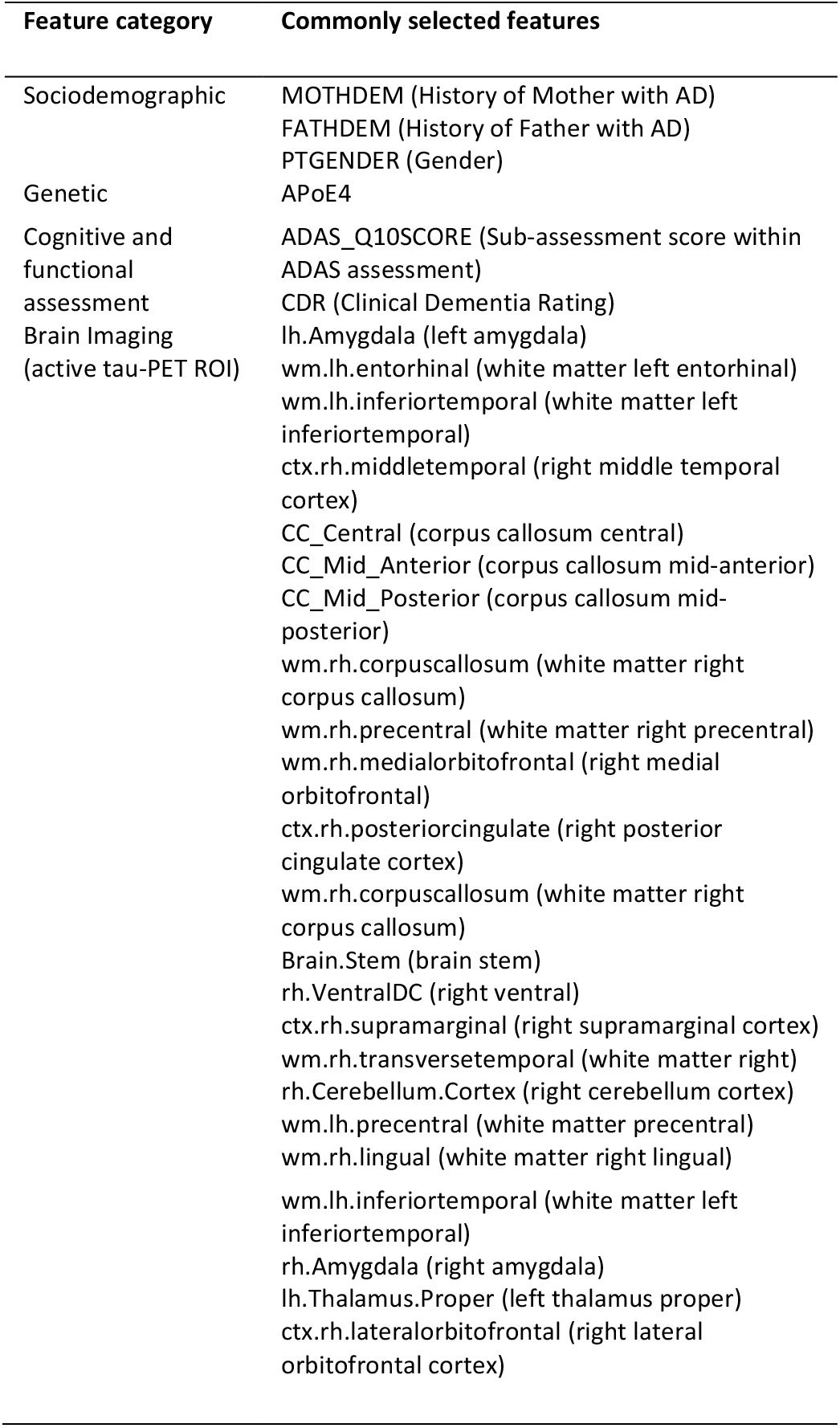
Common features selected from feature selection methods and denoising autoencoder

By quantifying the RMSE of the actual versus imputed value differences, Fig. 2 indicating the RMSE scores and imputation accuracy (ratio of number of correctly imputed values to the total number of imputed values), at different missingness levels of the 3 features. This indicated that our methods of implementing missing values and autoencoder-based imputation were conforming to expectation.

**Fig. 2.**
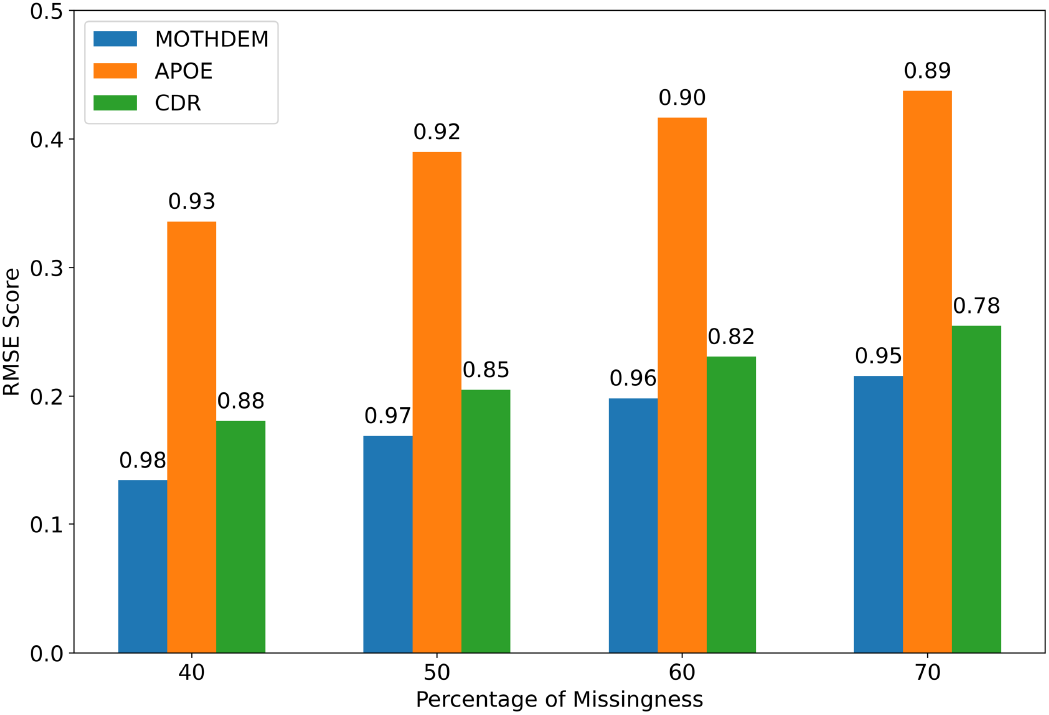
RMSE scores and imputation accuracy of the key features MOTHDEM, APoE4 and CDR after imputation.

### C. Robust AD classification of CN, MCI and AD of imputed data

After the imputation process, random forest classification with 10-fold cross validation was performed on the original dataset, imputed datasets of 40%, 50%, 60% and 70% missingness in MOTHDEM, CDR and APoE4. We also performed the same classification for dataset with only the common features selected using the three feature selection methods and included the important features that performed during each imputation process from the denoising autoencoder in it (see Supplementary Table 1 for details).

These results revealed the accuracy, precision, recall, and F1 score as 0.85, 0.85, 0.85, and 0.84, respectively, for the original dataset, which indicating a robust classification performance. These metrics underscore the efficacy of the random forest classifier in accurately predicting AD classes using the original dataset.

Subsequently the imputed datasets showed consistent classification accuracies, ranging from 0.71 to 0.83 across different missingness levels (Supplementary Table 1), indicating that the denoising autoencoder effectively imputed missing values without compromising classification performance. Additionally, classifiers trained on feature-selected datasets exhibited higher classification performance (0.89), suggesting that feature selection techniques retained relevant information for AD prediction, contributing to the classifier’s robustness.

We further evaluated the random forest classification’s precision, recall and F1 score of each of the 3 classes of the target variable AD_LABEL (with classes CN, MCI and AD). Fig. 3 summarises the results for each dataset highlighting the F1 scores of the three classes (see Supplementary Table 2 for detailed results, including precision and recall).

**Fig. 3.**
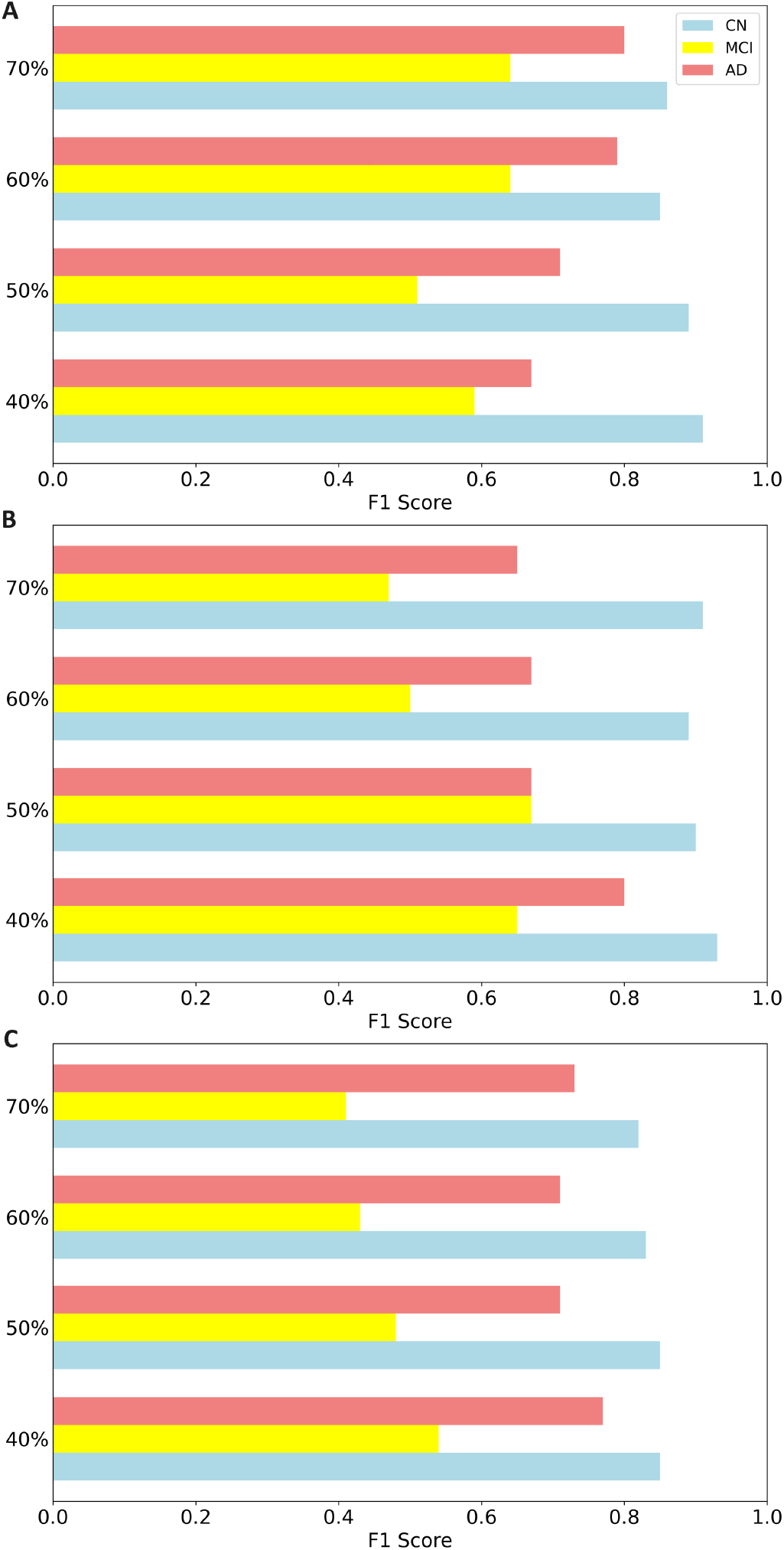
F1 scores after 3-class classification of the imputed datasets. (A) MOTHDEM (B) APoE (C) CDR.

Focusing on the F1 scores of each class (CN, MCI, and AD), we examined the classifier’s ability to accurately classify individuals with different disease progressive stages. In the original dataset, the random forest classifier demonstrated strong performance, with F1 scores of 0.91 for CN, 0.69 for MCI, and 0.85 for AD. This indicates robust classification across all progressive stages, which defining the effectiveness of the classifier on the complete dataset (see Supplementary Table 2 for further details). Noticeably, there was higher detriment to the F1 scores with missing CDR than MOTHDEM and APoE4.

## IV. DISCUSSION

Missing data is a common issue in healthcare datasets, and its presence can significantly impact the performance of predictive models, particularly in the context of Alzheimer’s disease diagnosis where accurate identification of relevant features is crucial. In our study, we addressed the challenge of missing data, specifically focusing on missing at random (MAR) patterns within the important features of the open ADNIMERGE-3 (ADNI) dataset. Previous studies did not consider such systematically selected features for data imputation evaluation. Specifically, we employed denoising autoencoder, a powerful nonlinear unsupervised learning technique, for imputing missing values in critical features such as demographics (MOTHDEM), genotype (APoE4), and CFA (CDR). By leveraging the underlying structure of the data, denoising autoencoder can effectively capture patterns and relationships, making it well-suited for imputing missing values in complex and comprehensive healthcare datasets like ADNI.

For the imputed datasets, particularly those with 40% missingness, exhibited robust performance in terms of recall, precision, and F1 score, suggesting that denoising autoencoder effectively imputed missing values without significantly compromising classification performance. In a previous study [16], they have achieved an accuracy of 78.67% in AD classification of the imputed AD dataset, but they did not specify the missing percentage of the imputation performed. In comparison, we achieved an overall accuracy of 79-85% in AD classification for our MAR datasets.

The random forest classifier used here for the three-class classification provided key insights into the performance of different datasets in identifying individuals with AD. The original dataset exhibited high F1 score (0.85) for the AD class, indicating its ability to effectively identify and accurately predict AD cases. Similarly, the feature-selected dataset demonstrated higher F1 score (0.89), suggesting that feature selection techniques retained crucial information for AD prediction. Interestingly, the performances were slightly higher than that for the original dataset, possibly due to reduction in model complexity with only the relevant features.

The F1 scores on the imputed datasets for each class across different levels of missingness provided valuable insights into the impact of imputation on classification performance. Generally, we observed a trend of decreasing F1 scores with increasing levels of missingness in the imputed datasets. In the case of MOTHDEM, the F1 scores for CN and MCI exhibited gradual decrease as the percentage of missingness increased, while the F1 score for AD remained relatively stable. This suggests that the imputation process may have a more pronounced effect on the classification of CN and MCI individuals compared to those with AD. Similarly, for APoE and CDR, we observed fluctuations in F1 scores across different classes with increasing levels of missingness. These variations highlight the importance of carefully considering the imputation strategy when dealing with heterogeneity and types of missing data, particularly in the context of AD classification.

The competitive performance by the selected feature dataset indicated that feature selection techniques retained relevant information essential for AD prediction. This highlights the importance of feature engineering and dimensionality reduction in enhancing predictive performance and reducing computational complexity. Furthermore, the inclusion of specific features identified in previous studies, such as APoE4, gender, (history of) mother with AD, active tau-PET in left amygdala, white matter in left entorhinal, white matter in left inferiortemporal ROIs, aligns with findings from a previous study using the same processed dataset [17], further validating the significance of these features in AD prediction.

There are several studies dealing with missing data imputation, including those using the ADNI dataset [11, 12, 13]. However, these studies did not apply deep learning methods and did not systematically investigate levels of missingness [6]. In comparison, our research utilised denoising autoencoder to impute missing data in diverse features of the ADNI dataset, accounted for various extreme levels of missingness and demonstrated robust performance in classification tasks. Hence, this study’s uniqueness lies in its integration of deep learning techniques with comprehensive handling of missing data, paving the way for more accurate diagnosis and understanding of AD progression.

While our study yielded valuable insights into the utilisation of denoising autoencoder for imputing missing values for AD diagnosis, there are notable limitations. Firstly, our analysis was confined to a single dataset, the comprehensive ADNIMERGE-3 dataset, and the generalisability of our findings to other datasets may be limited. Future studies should consider replicating our analysis on diverse datasets to assess the robustness and applicability of our approach across different cohorts and settings. Additionally, future studies could explore the integration of multiple imputation methods to enhance imputation accuracy and mitigate potential biases during imputation.

In conclusion, our study highlights the importance of using autoencoder for robust data imputation for high-performance machine classification across different disease progressive stages. Despite variations in missingness levels and feature selection strategies, the classifier consistently demonstrated strong performance metrics for predicting AD, underscoring its potential diagnostic utility in clinical decision-making and disease management.

## Data Availability

Source codes in the present study are available upon reasonable request to the authors. The original data used in the present study are available at Alzheimer's Disease Neuroimaging Initiative (ADNI).

https://adni.loni.usc.edu/

## ACKNOWLEDGEMENTS

Data collection and sharing for this project was funded by the Alzheimer’s Disease Neuroimaging Initiative (ADNI) (National Institutes of Health Grant U01 AG024904) and DOD ADNI (Department of Defense award number W81XWH-12-2-0012). ADNI is funded by the National Institute on Aging, the National Institute of Biomedical Imaging and Bioengineering, and through generous contributions from the following: AbbVie, Alzheimer’s Association; Alzheimer’s Drug Discovery Foundation; Araclon Biotech; BioClinica, Inc.; Biogen; Bristol-Myers Squibb Company; CereSpir, Inc.; Cogstate; Eisai Inc.; Elan Pharmaceuticals, Inc.; Eli Lilly and Company; EuroImmun; F. Hoffmann-La Roche Ltd and its affiliated company Genentech, Inc.; Fujirebio; GE Healthcare; IXICO Ltd.; Janssen Alzheimer Immunotherapy Research & Development, LLC.; Johnson & Johnson Pharmaceutical Research & Development LLC.; Lumosity; Lundbeck; Merck & Co., Inc.; Meso Scale Diagnostics, LLC.; NeuroRx Research; Neurotrack Technologies; Novartis Pharmaceuticals Corporation; Pfizer Inc.; Piramal Imaging; Servier; Takeda Pharmaceutical Company; and Transition Therapeutics. The Canadian Institutes of Health Research is providing funds to support ADNI clinical sites in Canada. Private sector contributions are facilitated by the Foundation for the National Institutes of Health (www.fnih.org). The grantee organization is the Northern California Institute for Research and Education, and the study is coordinated by the Alzheimer’s Therapeutic Research Institute at the University of Southern California. ADNI data are disseminated by the Laboratory for Neuro Imaging at the University of Southern California.

