## Supplementary Materials for "Autoencoder Imputation of Missing Heterogeneous Data for Alzheimer’s Disease Classification"

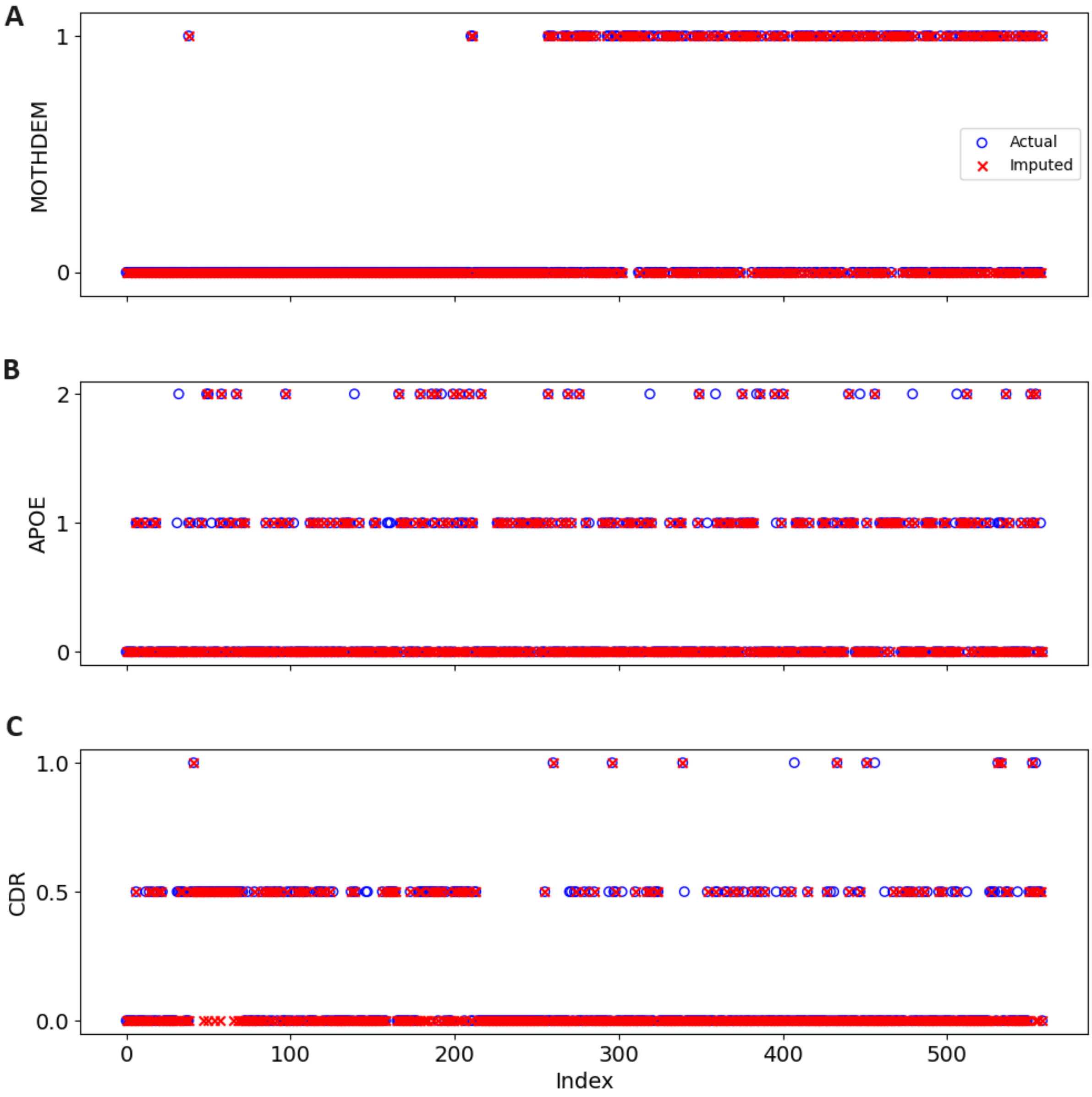


Supplementary Fig.1. Actual and imputed values for 3 key features MOTHDEM, APoE4 and CDR, each with 40% missingness. (A) History of mother with AD, MOTHDEM. Yes (1) or no (0). (B) Number of inherited APoE4 alleles. (C). CDR scores.

Supplementary Table 1. Classification performance for the original, feature selected, and imputed datasets. Bold: higher classification accuracy values.

| **Datasets** | **Accuracy** | **Precision** | **Recall** | **F1 Score** |
|---|---|---|---|---|
| Original | **0.85** | 0.85 | 0.85 | 0.84 |
| Features selected | **0.89** | 0.89 | 0.89 | 0.89 |
| 40% MOTHDEM (Missingness) | 0.80 | 0.80 | 0.80 | 0.80 |
| 50% MOTHDEM (Missingness) | 0.79 | 0.77 | 0.79 | 0.78 |
| 60% MOTHDEM (Missingness) | 0.78 | 0.79 | 0.78 | 0.78 |
| 70% MOTHDEM (Missingness) | 0.79 | 0.79 | 0.79 | 0.79 |
| 40% APoE4 (Missingness) | **0.83** | 0.83 | 0.83 | 0.83 |
| 50% APoE4 (Missingness) | 0.80 | 0.79 | 0.80 | 0.80 |
| 60%APoE4 (Missingness) | 0.78 | 0.77 | 0.78 | 0.77 |
| 70% APoE4 (Missingness) | 0.78 | 0.77 | 0.78 | 0.77 |
| 40% CDR (Missingness) | 0.76 | 0.77 | 0.76 | 0.76 |
| 50% CDR (Missingness) | 0.74 | 0.75 | 0.74 | 0.74 |
| 60% CDR (Missingness) | 0.72 | 0.72 | 0.72 | 0.72 |
| 70% CDR (Missingness) | 0.71 | 0.71 | 0.71 | 0.71 |

Supplementary Table 2. Classification performance for the original, feature selected and imputed datasets based on classes.

| **Datasets** | **Classes** | **Precision** | **Recall** | **F1 Score** |
|---|---|---|---|---|
| Original | 1 - CN | 0.90 | 0.92 | 0.91 |
| | 2 - MCI | 0.76 | 0.63 | 0.69 |
| | 3 - AD | **0.73** | **1.00** | 0.85 |
| Feature selected | 1 - CN | 0.91 | 0.91 | 0.91 |
| | 2 - MCI | 0.93 | 0.79 | 0.85 |
| | 3 - AD | **0.85** | **0.99** | 0.91 |
| 40% Mothdem (Missingness) | 1 - CN | 0.90 | 0.92 | 0.91 |
| | 2 - MCI | 0.62 | 0.56 | 0.59 |
| | 3 - AD | 0.62 | 0.71 | 0.67 |
| 50% Mothdem (Missingness) | 1 - CN | 0.87 | 0.92 | 0.89 |
| | 2 - MCI | 0.60 | 0.44 | 0.51 |
| | 3 - AD | 0.65 | 0.79 | 0.71 |
| 60% Mothdem (Missingness) | 1 - CN | 0.85 | 0.85 | 0.85 |
| | 2 - MCI | 0.60 | 0.68 | 0.64 |
| | 3 - AD | 0.88 | 0.71 | 0.79 |
| 70% Mothdem (Missingness) | 1 - CN | 0.83 | 0.90 | 0.86 |
| | 2 - MCI | 0.68 | 0.61 | 0.64 |
| | 3 - AD | 0.84 | 0.76 | 0.80 |
| 40% APoE4 (Missingness) | 1 - CN | 0.93 | 0.93 | 0.93 |
| | 2 - MCI | 0.68 | 0.63 | 0.65 |
| | 3 - AD | 0.62 | 0.71 | 0.67 |
| 50% APoE4 (Missingness) | 1 - CN | 0.89 | 0.92 | 0.90 |
| | 2 - MCI | 0.62 | 0.56 | 0.59 |
| | 3 - AD | 0.67 | 0.71 | 0.69 |
| 60% APoE4 (Missingness) | 1 - CN | 0.87 | 0.92 | 0.89 |
| | 2 - MCI | 0.57 | 0.44 | 0.50 |
| | 3 - AD | 0.62 | 0.71 | 0.67 |
| 70% APoE4 (Missingness) | 1 - CN | 0.86 | 0.96 | 0.91 |
| | 2 - MCI | 0.62 | 0.37 | 0.47 |
| | 3 - AD | 0.59 | 0.71 | 0.65 |
| 40% CDR (Missingness) | 1 - CN | 0.88 | 0.82 | 0.85 |
| | 2 - MCI | 0.52 | 0.56 | 0.54 |
| | 3 - AD | 0.71 | **0.86** | 0.77 |
| 50% CDR (Missingness) | 1 - CN | 0.87 | 0.83 | 0.85 |
| | 2 - MCI | 0.48 | 0.48 | 0.48 |
| | 3 - AD | 0.65 | 0.79 | 0.71 |
| 60% CDR (Missingness) | 1 - CN | 0.83 | 0.83 | 0.83 |
| | 2 - MCI | 0.46 | 0.41 | 0.43 |
| | 3 - AD | 0.65 | 0.79 | 0.71 |
| 70% CDR (Missingness) | 1 - CN | 0.82 | 0.82 | 0.82 |
| | 2 - MCI | 0.45 | 0.37 | 0.41 |
| | 3 - AD | 0.63 | 0.86 | 0.73 |
